# Clinical characteristics of COVID-19 infection in pregnant women: a systematic review and meta-analysis

**DOI:** 10.1101/2020.04.05.20053983

**Authors:** Sina Arabi, Golnaz Vaseghi, Zahra Heidari, Laleh Shariati, Bahareh Amin, Harunor Rashid, Shaghayegh Haghjooy Javanmard

## Abstract

**Background:** On December 2019, Novel coronavirus disease (COVID-19) was detected in Wuhan, China, and then spread around the world. There is little information about effects of COVID-19 on Pregnant women and newborns as a sensitive population. The current study is a systemic review and Meta-analysis to measure the risks and determine the presentations of COVID-19 in pregnant women and newborn.

**Methods:** online data bases were searched on march 20. Heterogeneity of the included studies was assessed using the Cochran Q test and Higgins I^2^ statistic and expressed as percentage. All data were analyzed with 95% confidence intervals.

**Results:** A total of 7 studies involving 50 participants with Positive test of COVID-19 were enrolled. Mean age of pregnant women was 30.57 years old and the Mean Gestational age was 36.9 weeks. Other variables such as Apgar score, birth weight, Sign and symptoms, Complications and Laboratory data were Analyzed.

**Conclusion:** Our findings showed same clinical characteristics in pregnant women as in non-pregnant adults, with the main symptoms being cough and fever. No vertical transmission was seen and all patients delivered healthy neonates. Our findings would be of great help to the decision making process, regarding the management of pregnant women diagnosed with COVID-19.

## Introduction

Severe acute respiratory syndrome coronavirus 2 (SARS-CoV-2) has spread rapidly since December 2019, in Wuhan, China, with an estimated mortality risk of ∼2% (1). Followed by SARS-CoV-2, an epidemic outburst in other regions of the country and worldwide is happening (2).

Coronaviruses mainly create enzootic infections in mammals and birds; lately, a capability to infect humans has been demonstrated as well (3). In less than seven days, the clinical symptoms begin, such as fever, fatigue, nasal congestion, cough, and other indications of upper respiratory tract involvements. Then, it can develop to a severe form of disease with severe chest symptoms and dyspnea, in about 75% of patients. In the day of 10 to 14 of a symptomatic infection, the pneumonia mostly occurs. Noticeable indications of viral pneumonia, such as blood gas deviations, and low oxygen saturation, leading to apparent alterations in chest X-rays, and finally lung abnormalities have been seen. Lymphopenia and elevation of inflammatory markers seems to be common (1, 4).

In cases of laboratory-confirmed SARS-CoV-2 infection, some studies described the medical, experimental, epidemiological and radiological specifications, as well as potential therapy and clinical end-results of patients (1, 5). However, no reliable evidence is as yet available to demonstrate clinical feature of COVID-19 infection in pregnant women.

The new coronavirus is genetically closer to SARS-CoV-1, and homology modelling has showed that it has a receptor binding domain structure like SARS-CoV-1, suggesting a comparable pathogenesis with SARS-CoV-1 infection (6). Recent studies have revealed that pregnant women with SARS-CoV-1 show high occurrence of adverse maternal and neonatal complications including preterm delivery, spontaneous abortion, intrauterine growth limitation, renal failure, endotracheal intubation usage and the intensive care unit admission and minimal chance of vertical transmission (7, 8).

Some investigations reported the medical manifestation and vertical transmission of COVID-19 in pregnancies (9-11). Because of the low power of study and small quantity of the samples, there are many essential questions that have to be addressed quickly, including whether severity of symptoms is the greater in pregnant women with COVID-19 as the sensitive population, if expecting mothers with COVID-19 die or give birth earlier, and finally whether COVID-19 could disseminate vertically and infect the embryo. Therefore, to better manage the infection of pregnant women, the present systematic review and meta-analysis has been launched to evaluate clinical manifestation of COVID-19 in the affected expecting women and assess its effect on the pregnancy end-results and neonatal well-being.

## Methods

### Search strategy and selection criteria

A systematic search was carried out in databases (PubMed, Embase and Cochrane Library) to identify published studies included epidemiological studies and case-control studies. the 2019 novel coronavirus (2019-nCoV), in accordance with the Preferred Reporting Items for Systematic Reviews and Meta-Analyses (PRISMA) guidelines. There were two independent reviewers 2019-nCoV, respectively. We used “2019 Novel coronavirus”, “Wuhan virus”, “covid-19” and “Pregnancy “, “gestation” to identify the relevant studies. For the 2019-nCoV, we searched for all studies published in all language between till 20 March 2020. We used English and Chinese studies.

### Statistical analysis

The meta-analysis was performed using “metan” and “metaprop” programs in STATA version 11 (STATA, College Station, TX, USA) based on clinical characteristics of COVID-19 infection as input. The meta-analysis resulted in pooled (overall) mean and prevalence of characteristics with a 95% confidence interval (95% CI). Heterogeneity of the included studies was assessed using the Cochran Q test and Higgins’ I^2^ statistic and expressed as percentage. Values of 25%, 50%, and 75% for I2 were considered as low, medium, and high levels of heterogeneity, respectively. Data were pooled using fixed or random effects model as appropriate. Publication bias was tested using Begg and Egger’s tests.

## Results

The schematic diagram of selection process and meta-analysis is shown in Figure 1. Seven studies with 50 participants have been included. Mean age of pregnant women included in this meta-analysis was 30.82 years old (95% CI: 28.54 – 33.10; Table 2 & Fig.2).

**Table 1.**
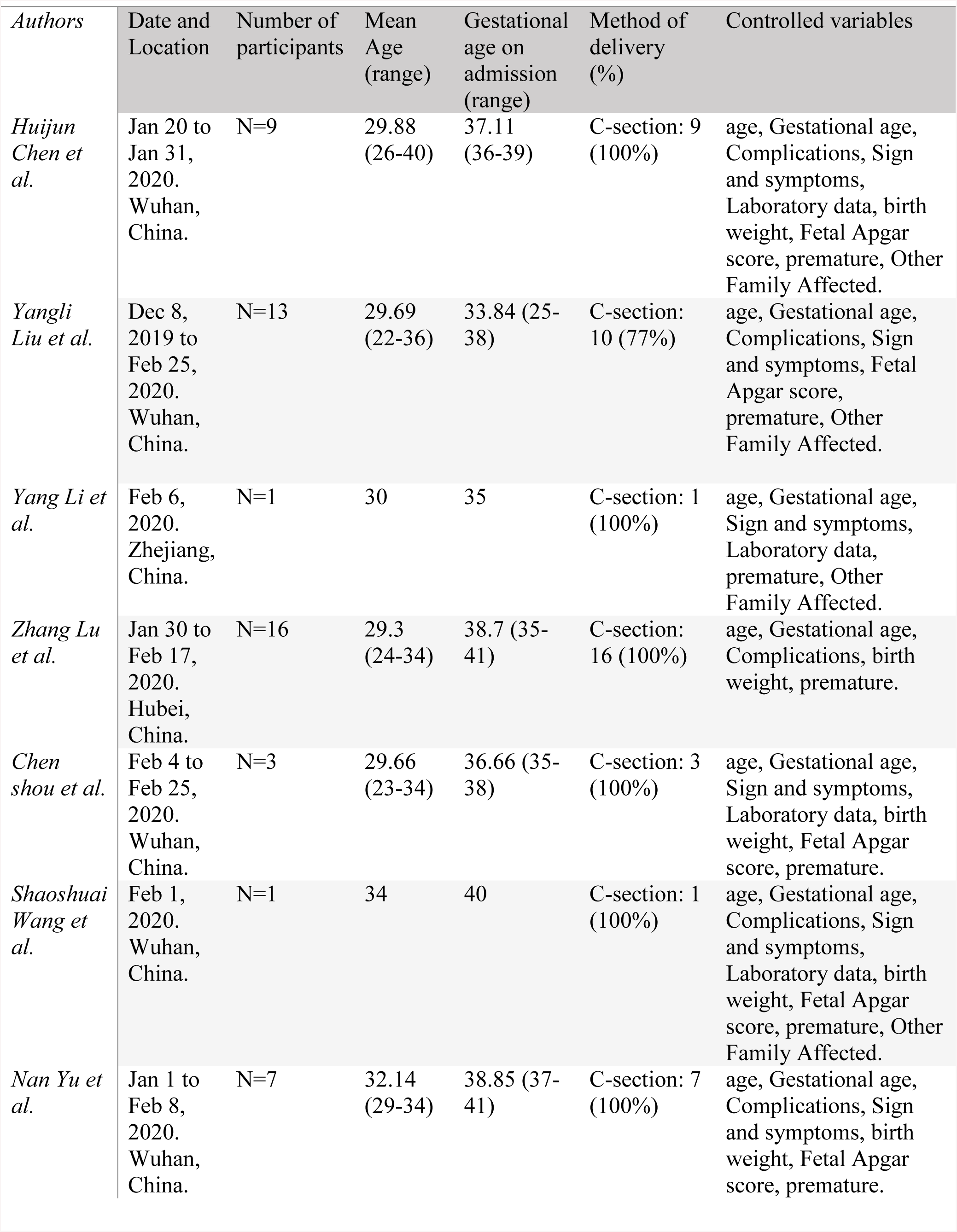
Included s*tudies, Characteristics Data and enrolled variables*.

**Table 2.** Pooled analysis of included Variables of studies.

|  | No. of studies | *Overall ES (95% CI) | Heterogeneity Tests |  | Publication bias p-values |  |
| --- | --- | --- | --- | --- | --- | --- |
|  |  |  | I<sup>2</sup> (%) | p-value | Begg's test | Egger's test |
| *Age* | 7 | 30.82 (28.54 – 33.10) | 100 | <0.001 | 0.543 | 1 |
| *Gestational Age* | 7 | 37.21 (34.54 – 39.87) | 100 | <0.001 | 0.649 | 1 |
| *WBC Count* | 4 | 9.51 (7.89 – 11.12) | 100 | <0.001 | 0.718 | 1 |
| *C Reactive Protein* | 4 | 16.83 (10.87 – 22.79) | 100 | <0.001 | 0.718 | 1 |
| *Birth Weight* | 4 | 3.2 (3.2 – 3.2) | 0 | 0.741 | 1 | 0.197 |
| *Apgar Score* | 5 | 9.43 (8.98 – 9.89) | 100 | <0.001 | 0.782 | 1 |
| *C section* | 7 | 1 (0.95 - 1) | 0 | 0.45 | 0.133 | 0.573 |
| *Premature Birth* | 7 | 0.20 (0.04 - 0.41) | 28.67 | 0.21 | 0.548 | 0.434 |
| *Death* | 7 | 0 (0 - 0.02) | 0 | 0.94 | 0.133 | 0.573 |
| *Other Family Affected* | 4 | 0.61 (0.32 - 0.87) | 4.66 | 0.37 | 0.279 | 0.783 |
| *Prom* | 5 | 0.08 (0 - 0.23) | 0 | 0.85 | 1 | 0.337 |
| *Fetal Distress* | 4 | 0.07 (0 - 0.21) | 0 | 0.68 | 1 | 0.749 |
| *Preeclampsia* | 6 | 0 (0 - 0.07) | 0 | 0.86 | 0.851 | 0.589 |
| *Asphyxia* | 6 | 0 (0 - 0.04) | 0 | 0.95 | 0.348 | 0.376 |
| *Fever* | 5 | 0.77 (0.57 - 0.93) | 0 | 0.62 | 1 | 0.852 |
| *Postpartum fever* | 4 | 0.02 (0 - 0.37) | 58.08 | 0.07 | 0.174 | 0.530 |
| *Myalgia* | 4 | 0.01 (0 - 0.25) | 51.08 | 0.11 | 0.734 | 0.939 |
| *Malaise* | 5 | 0 (0 - 0.09) | 0 | 0.45 | 0.327 | 0.811 |
| *Rigor* | 5 | 0 (0 - 0.02) | 0 | 0.98 | 1 | - |
| *Cough* | 5 | 0.20 (0.02 – 0.46) | 31.6 | 0.21 | 0.624 | 0.466 |
| *Dyspnea* | 5 | 0.10 (0 - 0.27) | 0 | 0.92 | 0.142 | 0.1 |
| *Sore throat* | 5 | 0.03 (0 - 0.17) | 0 | 0.71 | 0.624 | 0.512 |
| *Diarrhea* | 5 | 0 (0 - 0.11) | 0 | 0.65 | 0.624 | 0.905 |
| *Chest pain* | 5 | 0 (0 - 0.02) | 0 | 0.98 | 1 | - |
| *Fatigue* | 5 | 0 (0 - 0.08) | 0 | 0.92 | 0.624 | 0.362 |

\*Values are mean (95%CI) or proportion (95%CI)

**Figure 1.**
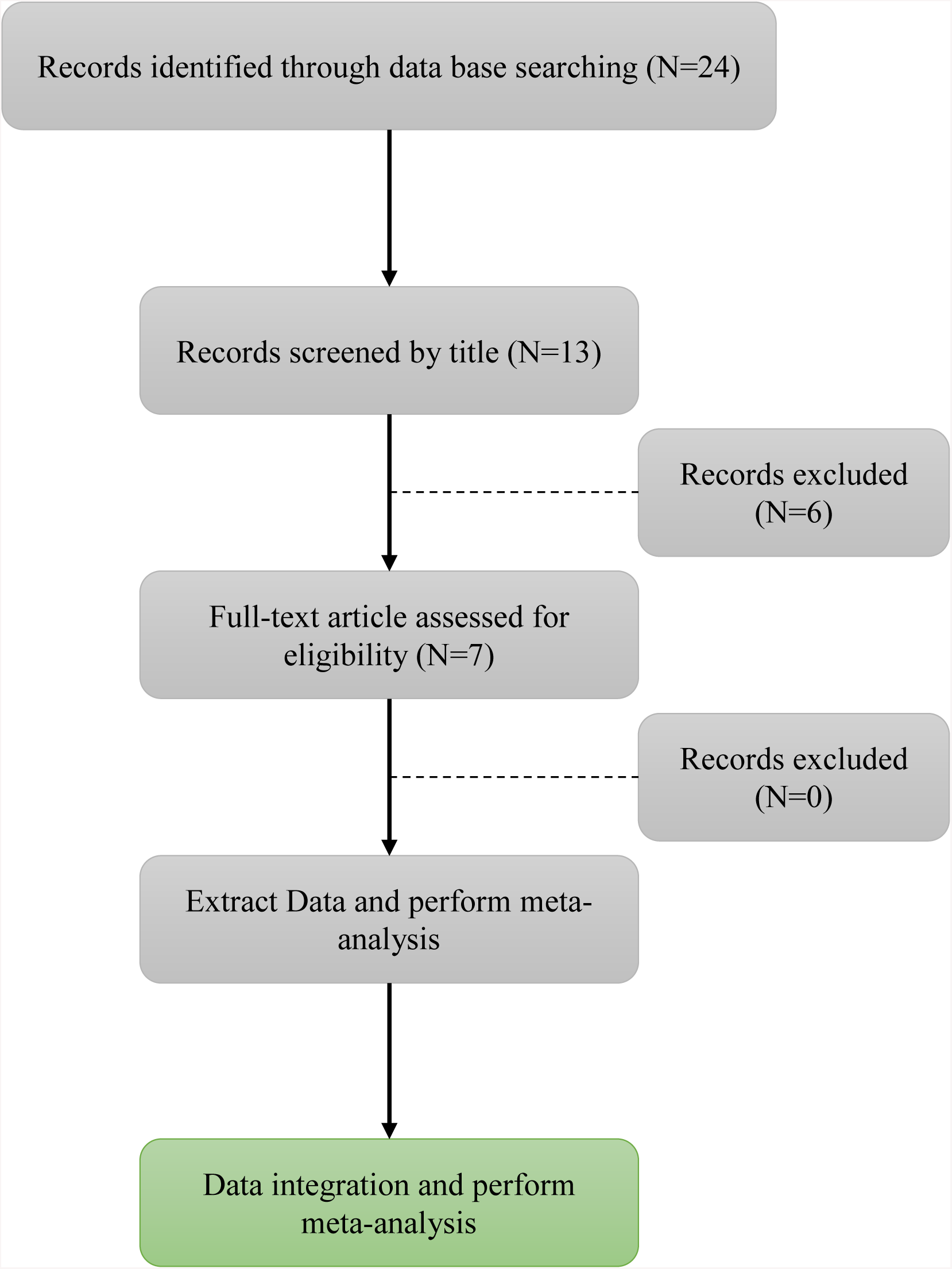
Schematic diagram of systemic review and selection process.

**Figure 2:**
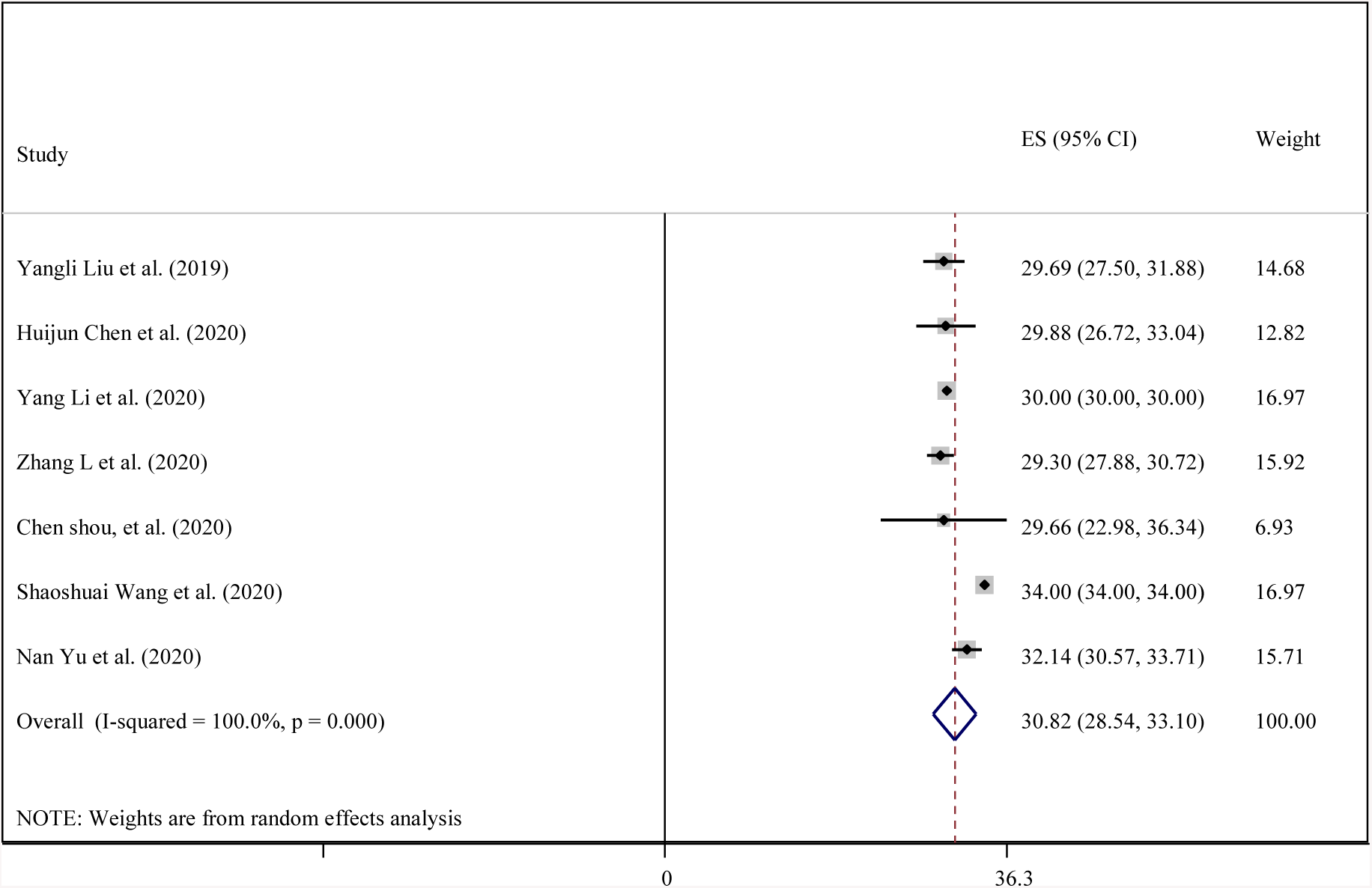
Forest plot showing Age status for individual studies included in a meta-analysis with 95% confidence intervals in pregnant women with COVID-19.

The pool mean of gestational age of pregnant women was 37.21 weeks, (95% CI: 34.54 – 39.87; I^2^= 100%; P-heterogeneity < 0.001; Table 2 & Fig.3), (Begg’s test P=0. 649, Egger’s test P=1).

**Figure 3:**
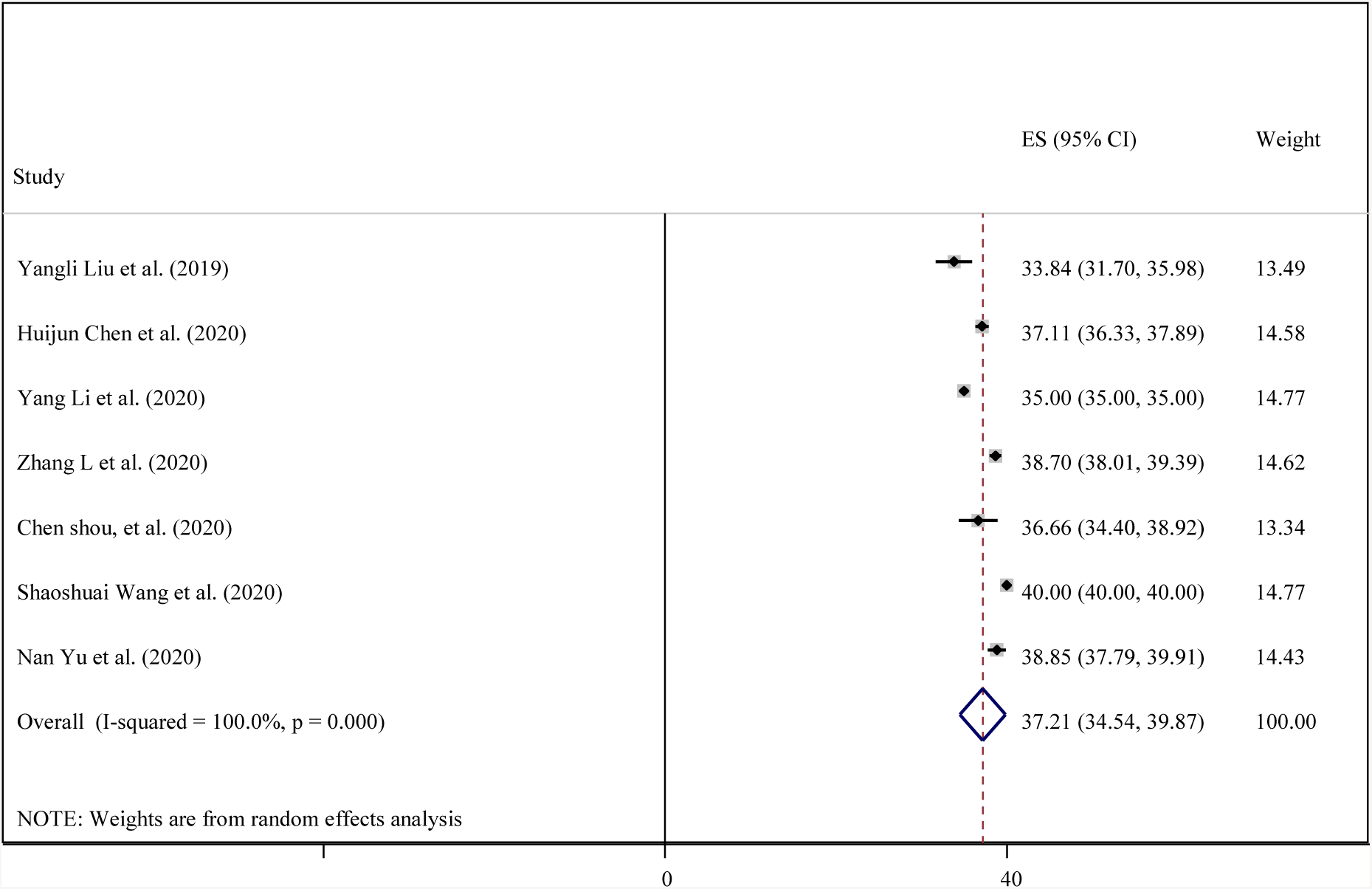
Forest plot showing Gestational Age status for individual studies included in a meta-analysis with 95% confidence intervals in pregnant women with COVID-19.

Four studies were combined to pool the mean of WBC count of pregnant women (9.51 × 10^9^ cells per dL, 95% CI: 7.89 – 11.12; I^2^= 100%; P-heterogeneity < 0.001; Table 2 & Fig.4). (Begg’s test P=0.718, Egger’s test P=1).

**Figure 4:**
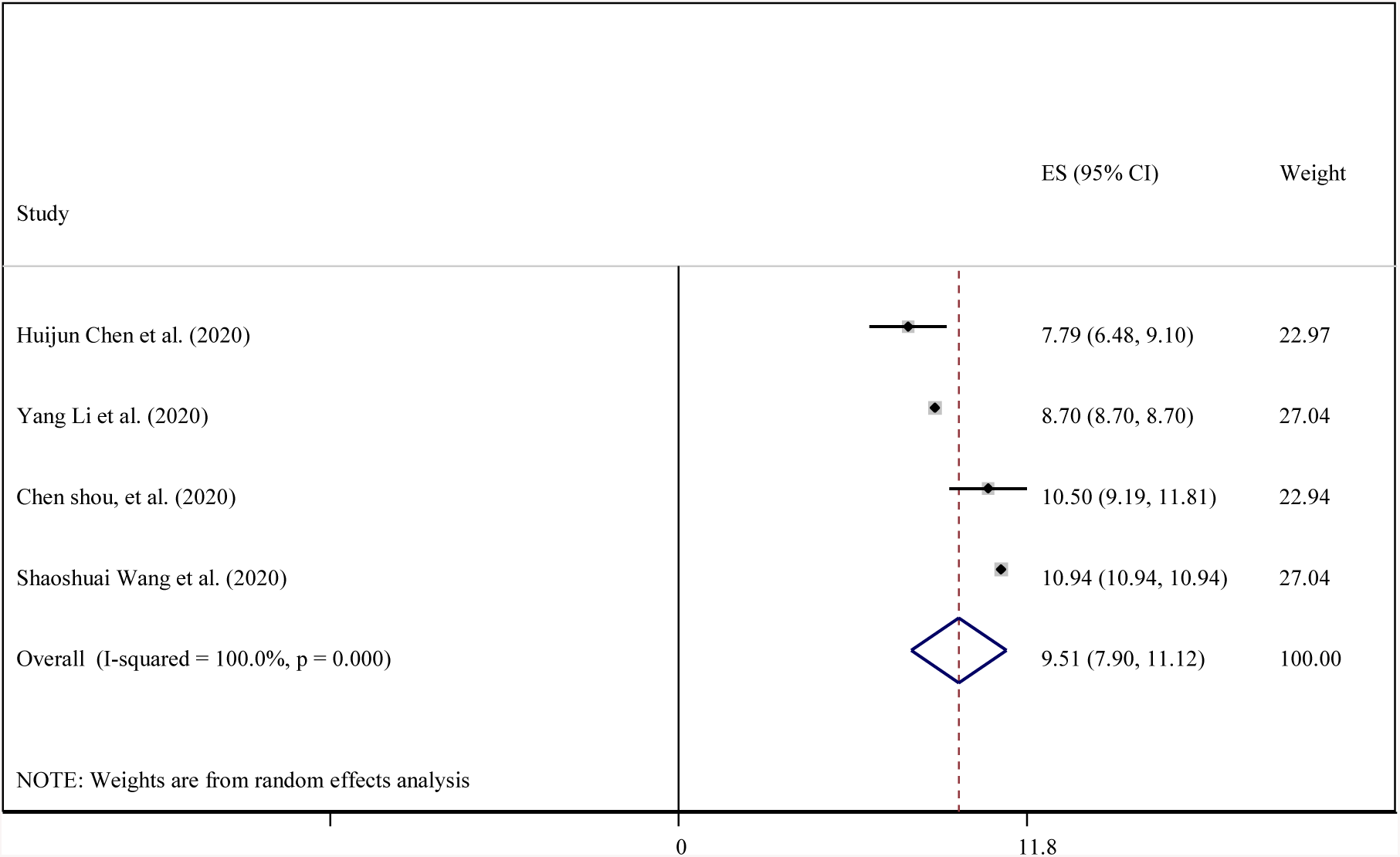
Forest plot showing WBC count for individual studies included in a meta-analysis with 95% confidence intervals in pregnant women with COVID-19.

the mean of C reactive protein of pregnant women was 16.83 mg/L, (95% CI: 10.87 – 22.79; I^2^= 100%; P-heterogeneity < 0.001; Table 2 & Fig.5),(Begg’s test P=0.718, Egger’s test P=1).

**Figure 5:**
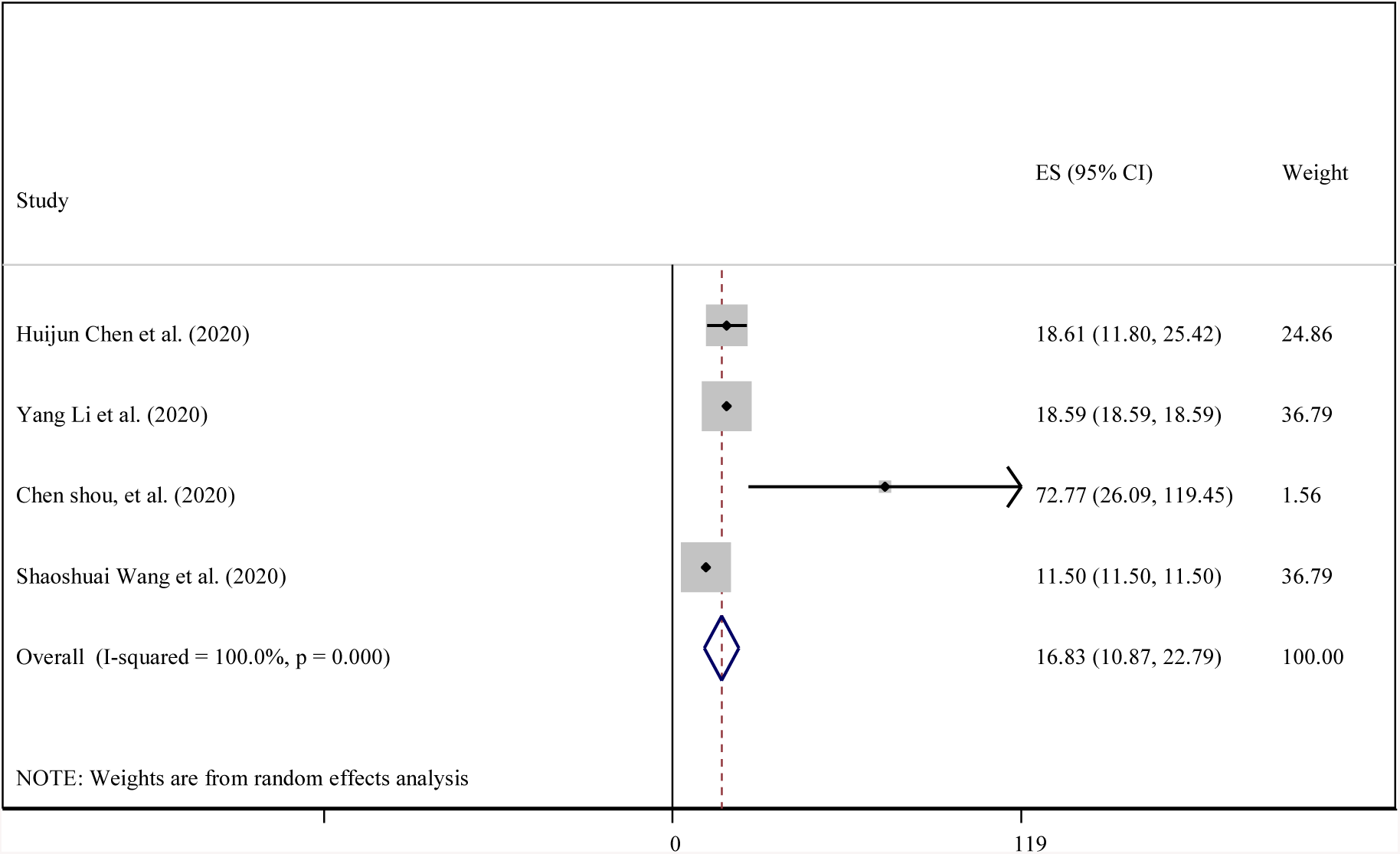
Forest plot showing C-reactive protein concentration for individual studies included in a meta-analysis with 95% confidence intervals in pregnant women with COVID-19.

Birth weight of pregnant women’s babies was 3.2 kg,(95% CI: 3.2 – 3.2; I^2^= 0%; P-heterogeneity = 0.741; Table 2 & Fig.6), (Begg’s test P=1, Egger’s test P=0.197).

**Figure 6:**
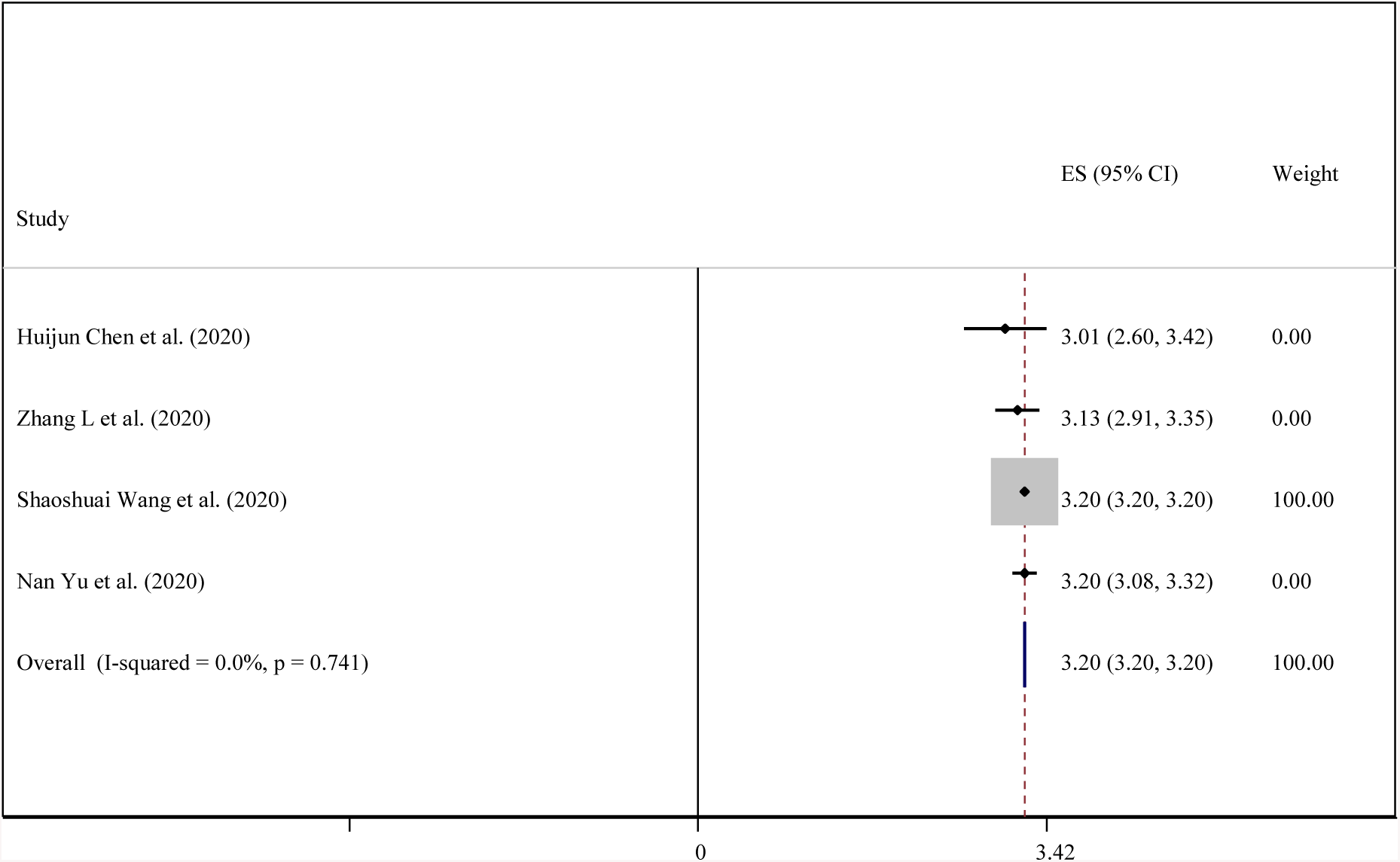
Forest plot showing Birth weight status for individual studies included in a meta-analysis with 95% confidence intervals in neonates from pregnant women with COVID-19.

Five studies were combined to pool the mean of Apgar score of infants, which was 9.43(95% CI: 8.98 – 9.89; I^2^= 100%; P-heterogeneity < 0.001; Table 2 & Fig.7). There was no significant study bias (Begg’s test P=0.782, Egger’s test P=1).

**Figure 7:**
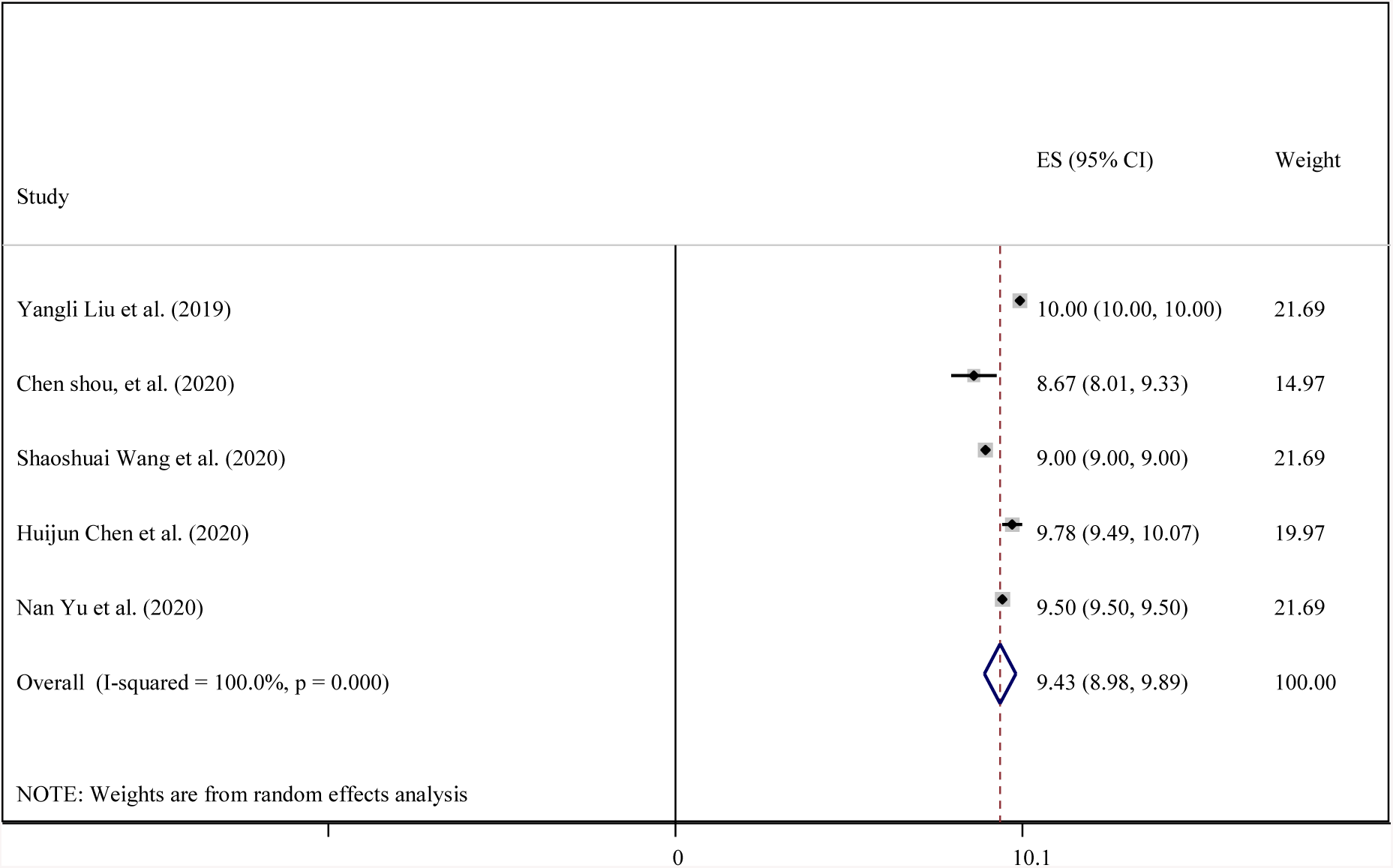
Forest plot showing Apgar-score status for individual studies included in a meta-analysis with 95% confidence intervals in neonates from pregnant women with COVID-19.

Fever had the highest overall prevalence among clinical characteristics of COVID-19 infection of pregnant women. We achieved an overall fever prevalence of 77% (95%-CI, 57–93%; I^2^= 0%; P-heterogeneity = 0.62; Table 2), (Begg’s test P=1, Egger’s test P=0.852). The cough symptom was in the next rank with prevalence of 20% (95%-CI; 2–46%; I^2^= 31.6%; P-heterogeneity = 0.21; Table 2). There was no significant study bias (Begg’s test P=0.624, Egger’s test P=0.466).

We achieved an overall premature prevalence of 20% (95%-CI, 4–41%; I^2^= 28.67%; P-heterogeneity = 0.21; Table 2 & Fig.9) through the meta-analysis with 7 studies included. There was no significant study bias (Begg’s test P=0.548, Egger’s test P=0.434).

**Figure 8:**
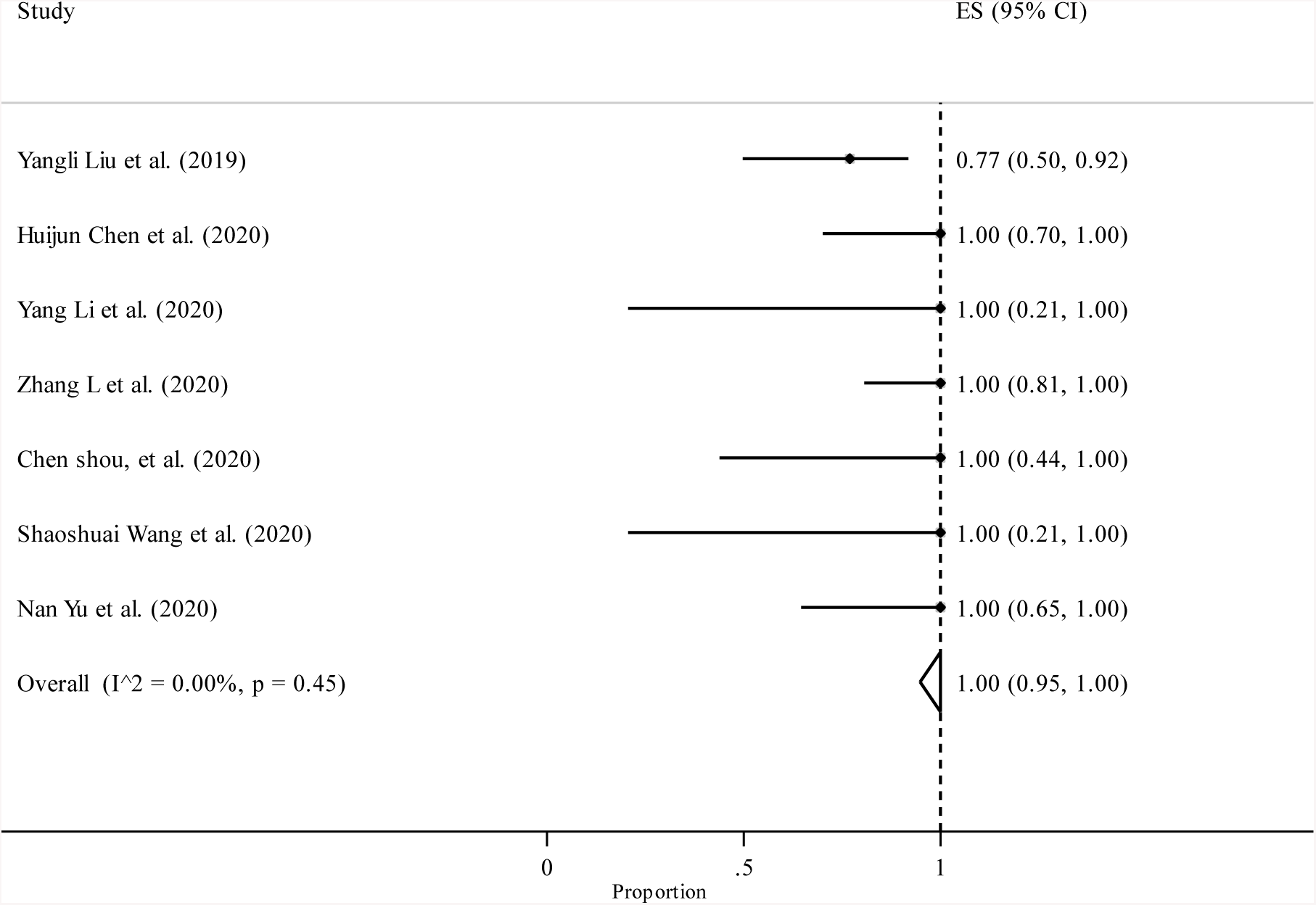
Forest plot showing C-section status for individual studies included in a meta-analysis with 95% confidence intervals in pregnant women with COVID-19.

**Figure 9:**
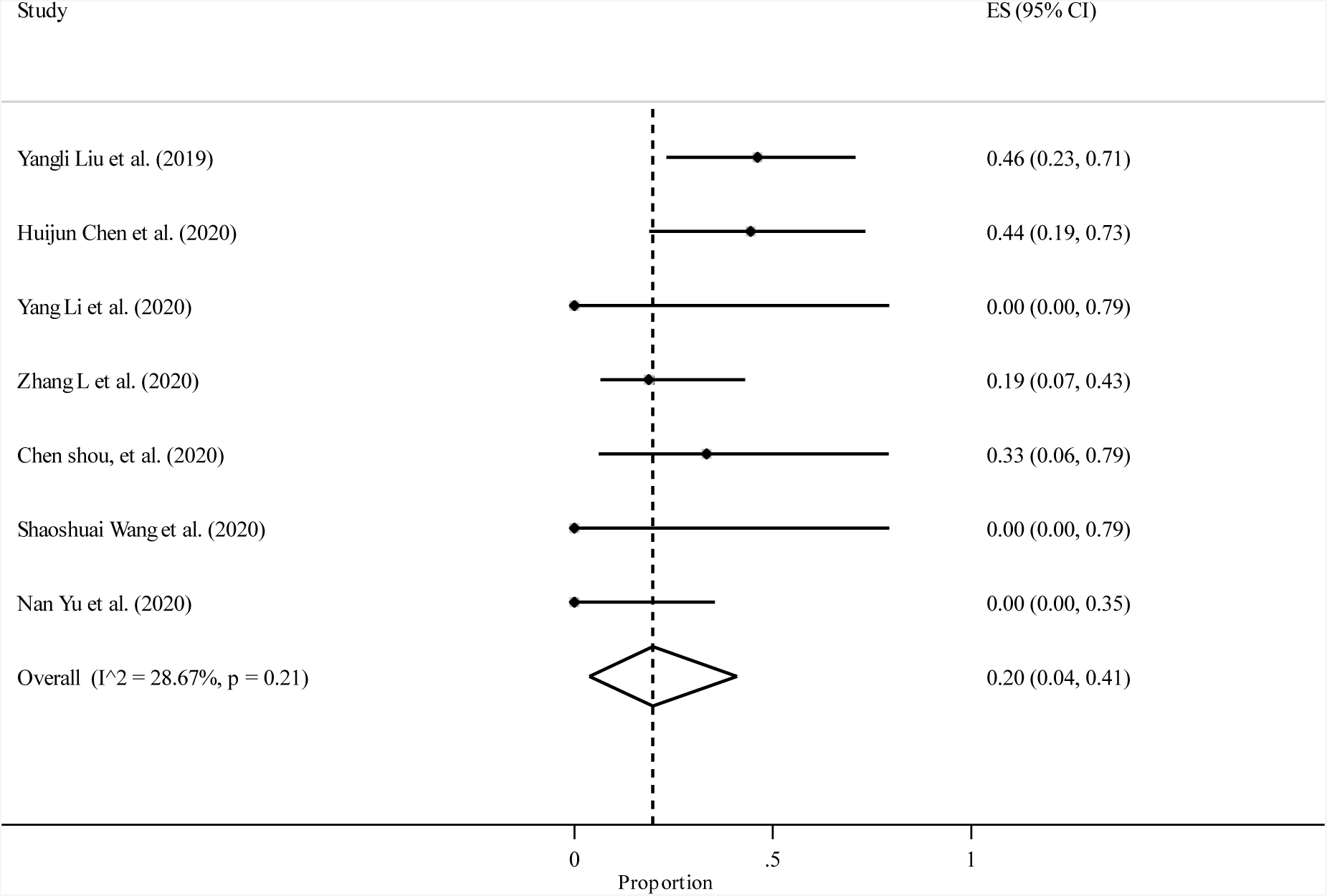
Forest plot showing prematurity status for individual studies included in a meta-analysis with 95% confidence intervals in neonates from pregnant women with COVID-19.

In this meta-analysis, we achieved an overall prevalence of more than 99% (95%-CI, 95–100%; I^2^= 0%; P-heterogeneity = 0.45; Table 2 & Fig.8) for cesarean section (C-section) through the combining 7 studies included. There was no significant study bias (Begg’s test P=0.133, Egger’s test P=0.573).

In this meta-analysis, we achieved an overall prevalence of 61% (95%-CI, 32–87%; I^2^= 4.66%; P-heterogeneity = 0.37; Table 2 & Fig.11) for affecting other family of participants through the combining 4 studies included. There was no significant study bias (Begg’s test P=0.279, Egger’s test P=0.783).

**Figure 10:**
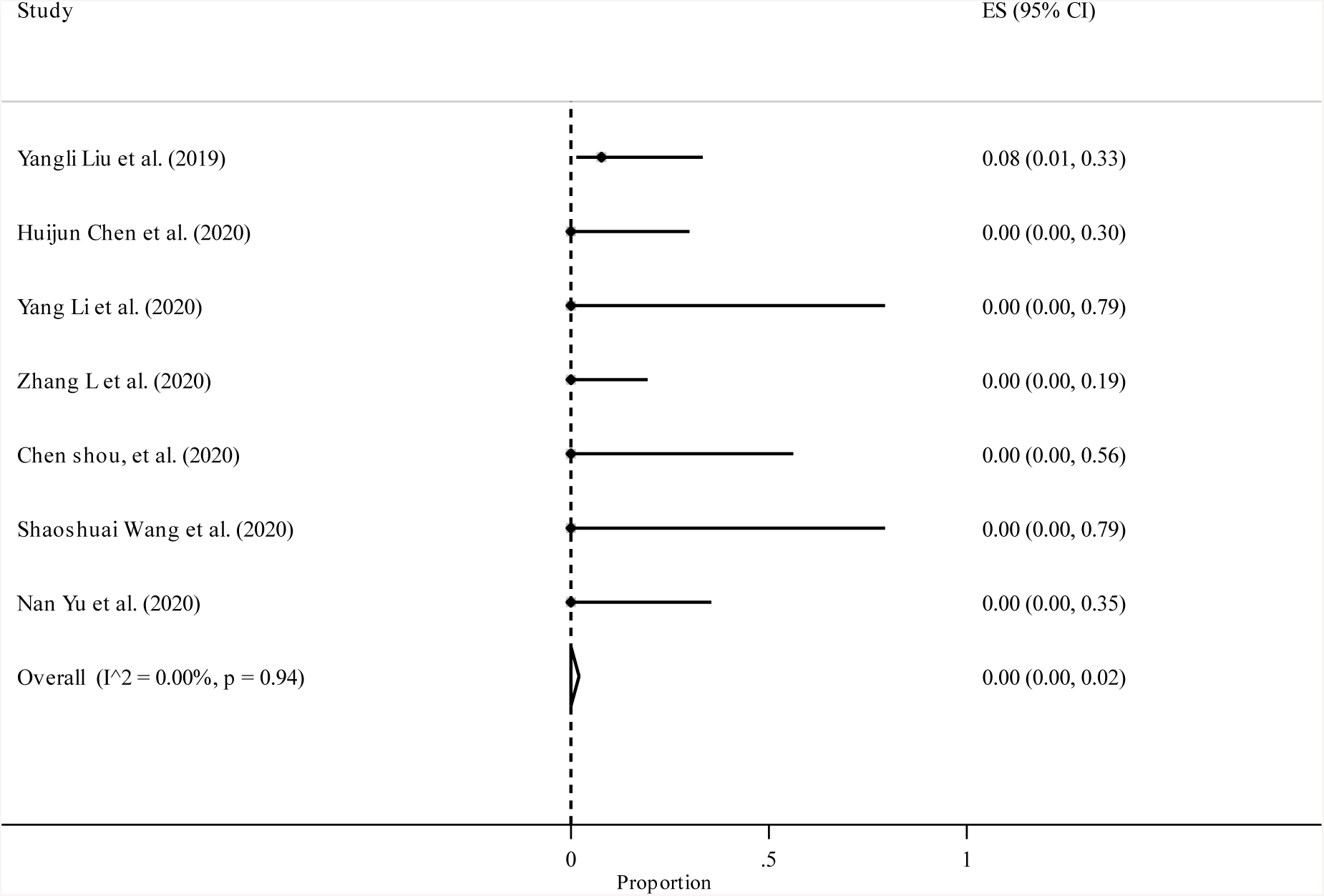
Forest plot showing death status for individual studies included in a meta-analysis with 95% confidence intervals in neonates from pregnant women with COVID-19.

**Figure 11:**
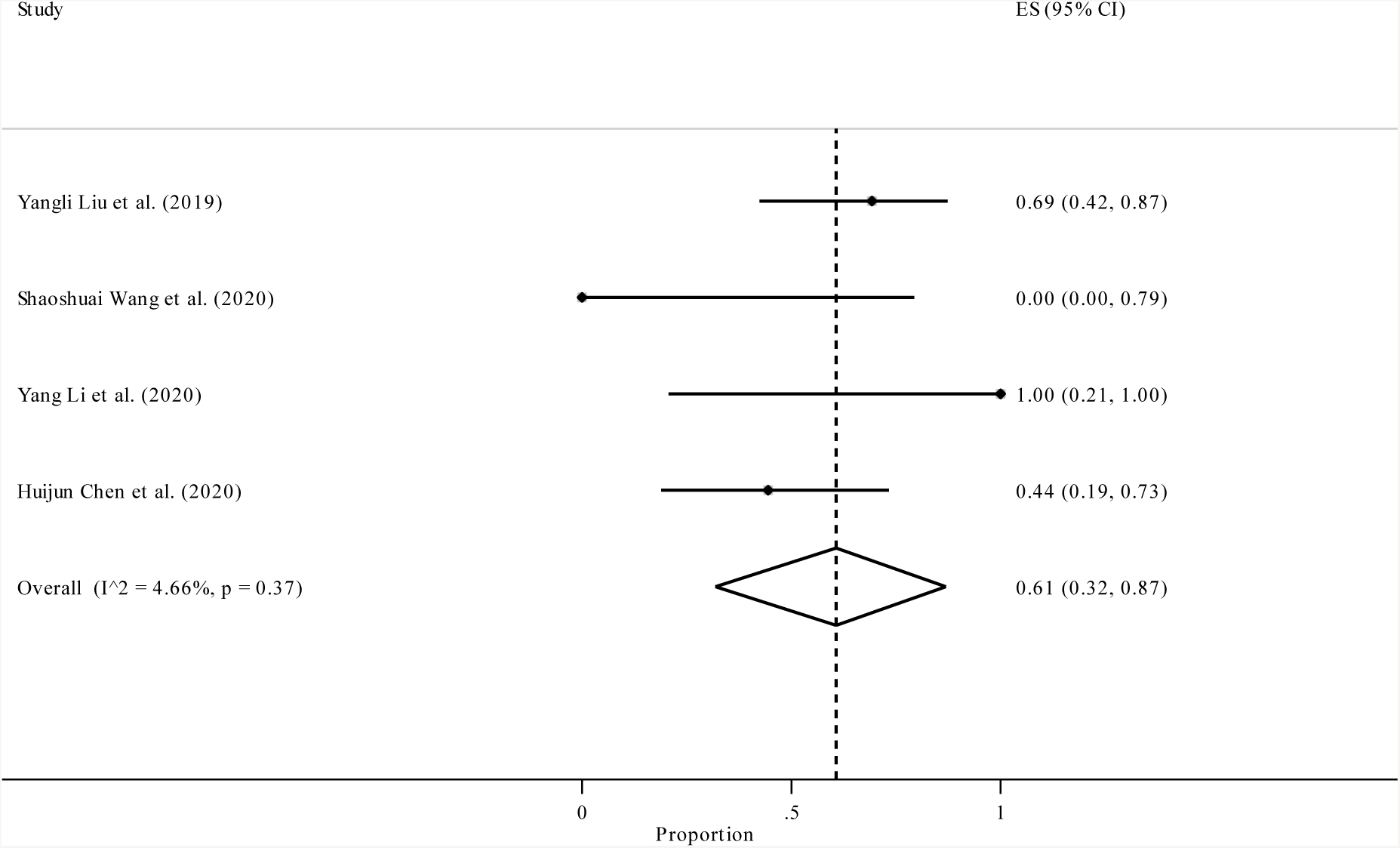
Forest plot showing other family affected status for individual studies included in a meta-analysis with 95% confidence intervals in pregnant women with COVID-19.

The current meta-analysis showed that the prevalence of death, preeclampsia, asphyxia, malaise, rigor, diarrhea, chest pain and fatigue is zero among pregnant women (Table 2). We achieved an overall prevalence of 8%, 7%, 2%, 1%, 10% and 3% for PROM (Prelabor rupture of membranes), fetal distress, postpartum fever, myalgia, dyspnea and sore throat, respectively (Table 2).

## Discussion

The emergence of the epidemic raised concerns regarding pregnant women as high-risk individuals. The prevention and management of COVID-19 in pregnant women and the possible risk of vertical transmission become the main concern (9). This meta-analysis is the first study to achieve a comprehensive pattern of the COVID-19 clinical features in pregnant women. We included seven studies with a total of 50 patients from China. The mean age of pregnant patients was 30-year old and all were in third trimesters at the time of manifestation.

The most pregnant patients with COVID-19 manifested mild to moderate signs, commonly fever (77 %) and cough (20%). The other symptoms prevalence and biochemical characterization are similar to non-pregnant patients with 2019-nCov infection (12). Respiratory viral diseases are indistinguishable from 2019-nCov infection due to nonspecific symptoms. To date, the laboratory tests have low specificity and cannot be used for definitive diagnosis of the COVID-19, and can only estimate the patient’s clinical condition, therefore PCR and radiological studies will be performed to confirm the disease (12).

This comprehensive study showed that the mortality rate is zero, although the sample size was limited. Also, there was no asphyxia, preeclampsia, rigor, malaise, chest pain, diarrhea and fatigue among pregnant women. However, differences in the quality of treatment in different countries (13) and virus genomic variation during spread (14) may cause these symptoms to show different rates elsewhere. In addition to the limiting number of studies included in this meta-analysis, another caveat is that all included studies were from China.

The results of four studies showed that 61% of the affected family members also had the disease. Because of close contact with the patient’s aerosols, the patient’s family members are at greater risk.

This meta-analysis showed that all pregnant women underwent C-section. It is not clear that the vertical transmission risk of 2019-nCov in C-section is lower than vaginal delivery (15). It seems that physicians think C-section is the better option for delivery. The surgery should be done in a negative-pressure operating room, and doctors should follow some protective measures like using a N95 mask, wearing a medical protective suit and goggles to avoid contamination with droplets from the surgical site (16).

Our analysis showed that all pregnant women delivered live and healthy infants that were negative tested for 2019-nCov. This result indicates that the possibility of vertical transmission is very low, and performing intra-operative hygiene procedures and transferring the baby to an isolated ward will help to protect the baby from getting infected with COVID-19.

## Conclusion

the clinical features of pregnant women and non-pregnant adult patients who developed COVID-19, are the same and the main symptoms were cough and fever among them. There is no evidence for vertical transmission in the third trimester of pregnancy, and all pregnant patients delivered healthy infants. Despite the limitations of this study, including the low number of studies included and being specific to China, regarding to emergency of the COVID-19 emergency, our findings would be of great help in making decision regarding to management and control of pregnant women with COVID-19.

## Data Availability

All data analysed during this study are included in this article.

## References

1. Huang C, Wang Y, Li X, Ren L, Zhao J, Hu Y, et al. Clinical features of patients infected with 2019 novel coronavirus in Wuhan, China. Lancet. 2020;395(10223):497–506.

2. Wu JT, Leung K, Leung GM. Nowcasting and forecasting the potential domestic and international spread of the 2019-nCoV outbreak originating in Wuhan, China: a modelling study. Lancet. 2020;395(10225):689–97.

3. Chan JF, Kok KH, Zhu Z, Chu H, To KK, Yuan S, et al. Genomic characterization of the 2019 novel human-pathogenic coronavirus isolated from a patient with atypical pneumonia after visiting Wuhan. Emerg Microbes Infect. 2020;9(1):221–36.

4. Zhang J, Zhou L, Yang Y, Peng W, Wang W, Chen X. Therapeutic and triage strategies for 2019 novel coronavirus disease in fever clinics. Lancet Respir Med. 2020;8(3):e11–e2.

5. Bastola A, Sah R, Rodriguez-Morales AJ, Lal BK, Jha R, Ojha HC, et al. The first 2019 novel coronavirus case in Nepal. Lancet Infect Dis. 2020;20(3):279–80.

6. Lu R, Zhao X, Li J, Niu P, Yang B, Wu H, et al. Genomic characterisation and epidemiology of 2019 novel coronavirus: implications for virus origins and receptor binding. Lancet. 2020;395(10224):565–74.

7. Lam CM, Wong SF, Leung TN, Chow KM, Yu WC, Wong TY, et al. A case-controlled study comparing clinical course and outcomes of pregnant and non-pregnant women with severe acute respiratory syndrome. BJOG. 2004;111(8):771–4.

8. Wong SF, Chow KM, Leung TN, Ng WF, Ng TK, Shek CC, et al. Pregnancy and perinatal outcomes of women with severe acute respiratory syndrome. Am J Obstet Gynecol. 2004;191(1):292–7.

9. Liu Y, Chen H, Tang K, Guo Y. Clinical manifestations and outcome of SARS-CoV-2 infection during pregnancy. J Infect. 2020.

10. Mardani M, Pourkaveh B. A Controversial Debate: Vertical Transmission of COVID-19in Pregnancy. Arch Clin Inect Dis. 2020;15(1):e102286.

11. Chen H, Guo J, Wang C, Luo F, Yu X, Zhang W, et al. Clinical characteristics and intrauterine vertical transmission potential of COVID-19 infection in nine pregnant women: a retrospective review of medical records. Lancet. 2020;395(10226):809–15.

12. Nascimento I, Cacic N, Abdulazeem H, Esfahani M, Jayarajah U, Jeroncic A, et al. Novel coronavirus infection in humans: a scoping review and meta-analysis. The Lancet. 2020.

13. Ji Y, Ma Z, Peppelenbosch M, Pan Q. Potential association between COVID-19 mortality and health-care resource availability. The Lancet Global Health. 2020;8(4):480.

14. Cao Y, Li L, Feng Z, Wan S, Huang P, Sun X, et al. Comparative genetic analysis of the novel coronavirus (2019-nCoV/SARS-CoV-2) receptor ACE2 in different populations. Cell Discov. 2020;6:11.

15. Li Y, Zhao R, Zheng S, Chen X, Wang J, Sheng X, et al. Lack of Vertical Transmission of Severe Acute Respiratory Syndrome Coronavirus 2, China. Emerg Infect Dis. 2020;26(6).

16. Xu X, Chen P, Wang J, Feng J, Zhou H, Li X, et al. Evolution of the novel coronavirus from the ongoing Wuhan outbreak and modeling of its spike protein for risk of human transmission. Sci China Life Sci. 2020;63(3):457–60.

